# A multiplex protein panel assay determines disease severity and is prognostic about outcome in COVID-19 patients

**DOI:** 10.1101/2021.12.03.21267253

**Authors:** Ziyue Wang, Adam Cryar, Oliver Lemke, Daniela Ludwig, Pinkus Tober-Lau, Elisa Theresa Helbig, Daniel Blake, Catherine S Lane, Rebekah L Sayers, Christoph Mueller, Johannes Zeiser, StJohn Townsend, Vadim Demichev, Michael Mülleder, Florian Kurth, Ernestas Sirka, Johannes Hartl, Markus Ralser

## Abstract

Global healthcare systems continue to be challenged by the COVID-19 pandemic, and there is a need for clinical assays that can both help to optimize resource allocation and accelerate the development and evaluation of new therapies. Here, we present a multiplex proteomic panel assay for the assessment of disease severity and outcome prediction in COVID-19. The assay quantifies 50 peptides derived from 30 COVID-19 severity markers in a single measurement using analytical flow rate liquid chromatography and multiple reaction monitoring (LC-MRM), on equipment that is broadly available in routine and regulated analytical laboratories. We demonstrate accurate classification of COVID-19 severity in patients from two cohorts. Furthermore, the assay outperforms established risk assessments such as SOFA and APACHE II in predicting survival in a longitudinal COVID-19 cohort. The prognostic value implies its use for support of clinical decisions in settings with overstrained healthcare resources e.g. to optimally allocate resources to severely ill individuals with high chance of survival. It can furthermore be helpful for monitoring of novel therapies in clinical trials.

## Introduction

COVID-19 continues to challenge healthcare systems worldwide despite vaccination efforts and novel treatments. This is particularly apparent in areas with limited vaccine uptake or supply. Currently, the outlook remains uncertain even in countries with high vaccination rates as the immunity conferred by the vaccines appears to diminish over time [1–5] and SARS-COV-2 variants with varying capacity to evade vaccine induced immunity continue to emerge [1,6–8]. As of today, novel variants of concern, such as B.1.1.529 (Omicron), are still on the rise, and may affect global medical care rapidly and in an unpredictable way [9].

Biomarker tests that can classify patients’ disease severity, and those that are prognostic, could help to mitigate the impact of critical treatment choice, by allowing to optimise resource allocation, specifically during a pandemic [10–12]. Indeed, clinical manifestation of COVID-19 is highly variable, which does create challenges in timely clinical decision making. For instance, ’happy hypoxia’ describes situations where COVID-19 patients report minor impairments, while molecular indicators such as blood oxygen levels, indicate they are, in fact, severely ill [13]. Furthermore, in situations when healthcare systems reach maximum capacity, prognostic tests could support difficult clinical decisions, for instance, to navigate through triaging situations, and to ensure that individuals with a high chance of survival are provided with a maximum available support, irrespective of age and comorbidities [14]. Prognostic and disease severity tests could further help to increase the likelihood of success and accelerate clinical trials, by improving the assessment of treatment efficacy of COVID-19 therapies or stratify patient populations that are to be included in the trials. Indeed, COVID-19 prompted the rapid repurposing and development of new treatments [15]. However, during a pandemic, there is a need to conduct trials in a timely manner, but specifically in ICU settings, study cohorts are often limited in size. When underpowered, clinical trials can lead to wrong decisions, i.e. if they lead to false positive and false negative assessments of a drug’s efficacy [16,17]. Disease severity and prognostic tests could hence help to extrapolate more and patient-specific information in early stage clinical trials. Unfortunately however, the reliability of several risk assessment scores conventionally used in ICU settings, such as the Acute Physiology And Chronic Health Evaluation (APACHE II), Charlson Comorbidity Index (CCI) and Sequential Organ Failure Assessment (SOFA) scores, appear to be limited for COVID-19 cases [18]. Combinations of generic clinical readouts, e.g. blood oxygen saturation, interleukin-6 concentration, have been considered for COVID-19 outcome prediction at various disease severity stages [19]. However, also here, several predictive models based on clinical parameters and routine tests were reported early in the pandemic, are now considered to be vulnerable to bias and might not be suitable for the clinic [20,21].

Protein biomarker signatures derived from plasma present a promising new alternative to obtain patient risk assessments in addition to the established parameters. Proteomic datasets have repeatedly been successful at classifying and predicting COVID-19 severity and outcome [10,11,18,22–24]. Indeed, specifically in severe COVID-19 cases, proteomic predictors routinely outperform the Apache II, CCI and SOFA scores [12,14,25,26]. In parallel, especially early in the pandemic, proteomics accelerated the characterisation of the antiviral host response, which greatly improved our understanding of the COVID-19 disease within a short time, by attributing the complement cascade, the coagulation system, and apoprotein function, to differences in COVID-19 pathology [10,11,18,22–24,26]. The application of discovery proteomic platforms in the clinical routine is, however, limited for technical, economic and regulatory reasons. With only recent exceptions [17,25], most discovery proteomics platforms make use of low flow rate chromatography, which provides high sensitivity, but requires a high degree of expert knowledge to reach a level of stability and throughput required in many routine laboratories [27]. Other limiting factors include the high amount of data processing, and stochastic elements that accompany data-dependent acquisition techniques that are dominating the proteomic landscape. Most discovery proteomics platforms cannot be accredited to existing regulatory standards either.

Leveraging the predictive value of the proteome in COVID-19, the objective of this study was to develop a COVID-19 biomarker panel assay which runs on proteomics platforms that could be deployed for clinical use within existing regulatory frameworks and on broadly available analytical instruments. Triple quadrupole mass spectrometers coupled to high flow liquid chromatography were the optimal choice for rapid test development and deployment as they are used in the clinic in other areas, for instance rare diseases, newborn screening and steroid hormone analysis [28–31]. Further, they are widely available in large hospital laboratories, diagnostic laboratories, regulated (e.g. CLIA) laboratories, and contract research organizations. Biomarker tests developed on this platform can be accredited to existing regulatory standards in GCP, ISO:17025, ISO:15189 and CLIA environments, standardised and transferred across different instruments, manufacturers and laboratories, and thus deployed at scale rapidly. Triple quadrupole mass spectrometry-based tests are cost effective to run at scale as the sample preparation can be automated, consumables costs for the MS runs are typically <£10 per test and the instrument uptime is typically >95%. The tests also integrate with existing workflows at clinical and analytical laboratories.

To bridge the gap from discovery proteomics to clinical application in COVID-19, we have mined discovery proteomics data from COVID-19 patients, and selected biomarkers that are informative about COVID-19 disease progression. The biomarkers were chosen for i) being differentially concentrated in plasma depending on the treatment escalation level, used as a measure of disease severity, ii) are up in patients at risk of aggravation, and iii) participating in biological processes that contribute to COVID-19 pathology (Supplementary Table 1). The COVID-19 severity biomarkers chosen include proteins that function in inflammation (e.g. C-reactive protein), coagulation and vascular dysfunction (e.g. von Willebrand factor), complement cascade (e.g. Complement C1q subcomponent subunit C) and diverse biological processes detected to be altered by COVID-19 (e.g. Cystatin C, a known marker of kidney dysfunction). We then established a targeted proteomics assay utilising multiple reaction monitoring (MRM) data acquisition mode on routinely used LC-MS/MS instrumentation. Employing calibration curves with synthetic reference and stable isotope labelled (SIL) internal standards, the assay provides potential for absolute quantification of up to 50 surrogate tryptic peptides corresponding to 30 plasma proteins. We have analytically validated the assay, and implemented it in two analytical laboratories employing two triple quadrupole LC-MS/MS platforms from different manufacturers. The assay was then applied to two cohorts to demonstrate that the presented MRM assay captures host response to SARS-CoV-2 and thereby classifies and predicts COVID-19 disease severity. In a longitudinal cohort, we tested the prognostic value of the severity panel. We found that the biomarker panel is predictive about survival weeks before outcome, and outperforms commonly used ICU risk assessment scores. The assay could hence expand the arsenal of existing risk assessment procedures with a COVID-19 associated test, to identify severely ill patients with a good prognosis, and direct them to the optimal treatment level.

## Results

### Peptide selection

50 peptides that corresponded to 30 plasma proteins (Supplementary Table 1) were selected from shotgun plasma proteomics data recorded on a deeply phenotyped COVID-19 patient cohort treated at Charité Universitätsmedizin Berlin (PA-COVID-19 study cohort, N=139 inpatients, for which 687 plasma proteomes were measured in time series) [14,32]. To create a panel for COVID-19 severity prediction, we selected peptides that change in abundance depending on the COVID-19 inpatient treatment escalation level, expressed on the WHO ordinal scale [33]. Further, the selected peptides are derived from proteins whose functions are associated with the COVID-19 host response, in particular the inflammatory and innate immune response, the complement cascade and coagulation (Fig. 2a). A third selection criterion was that an abundance change in the putative marker peptides were predictive about the future worsening of COVID-19 inpatients who were admitted with a milder disease which then deteriorated, and about the remaining time in hospital for COVID-19 inpatients, which in turn serves as a treatment-insensitive proxy for COVID-19 severity (Fig. 2 a, b) [14,18,22,25,34,35]. Finally, we tested if native reference peptides were unique to a corresponding protein within the human proteome in a Uniprot BLAST and manual human proteome FASTA text file search. This was true for 44/50 peptides. One (1) peptide returned no search results and the remaining 5 peptides (CQSWSSMTPHR, EITALAPSTMK, WEMPFDPQDTHQSR, DSGSYFCR, ASDTAMYYCAR) were shared across closely related protein isoforms or proteins with similar functions (Supplementary Table 1). All 50 peptides were included in the panel composition as they fulfilled the selection criteria described above. SIL internal standards (ISTDs) were designed based on the selected native peptides.

**Figure 1.**
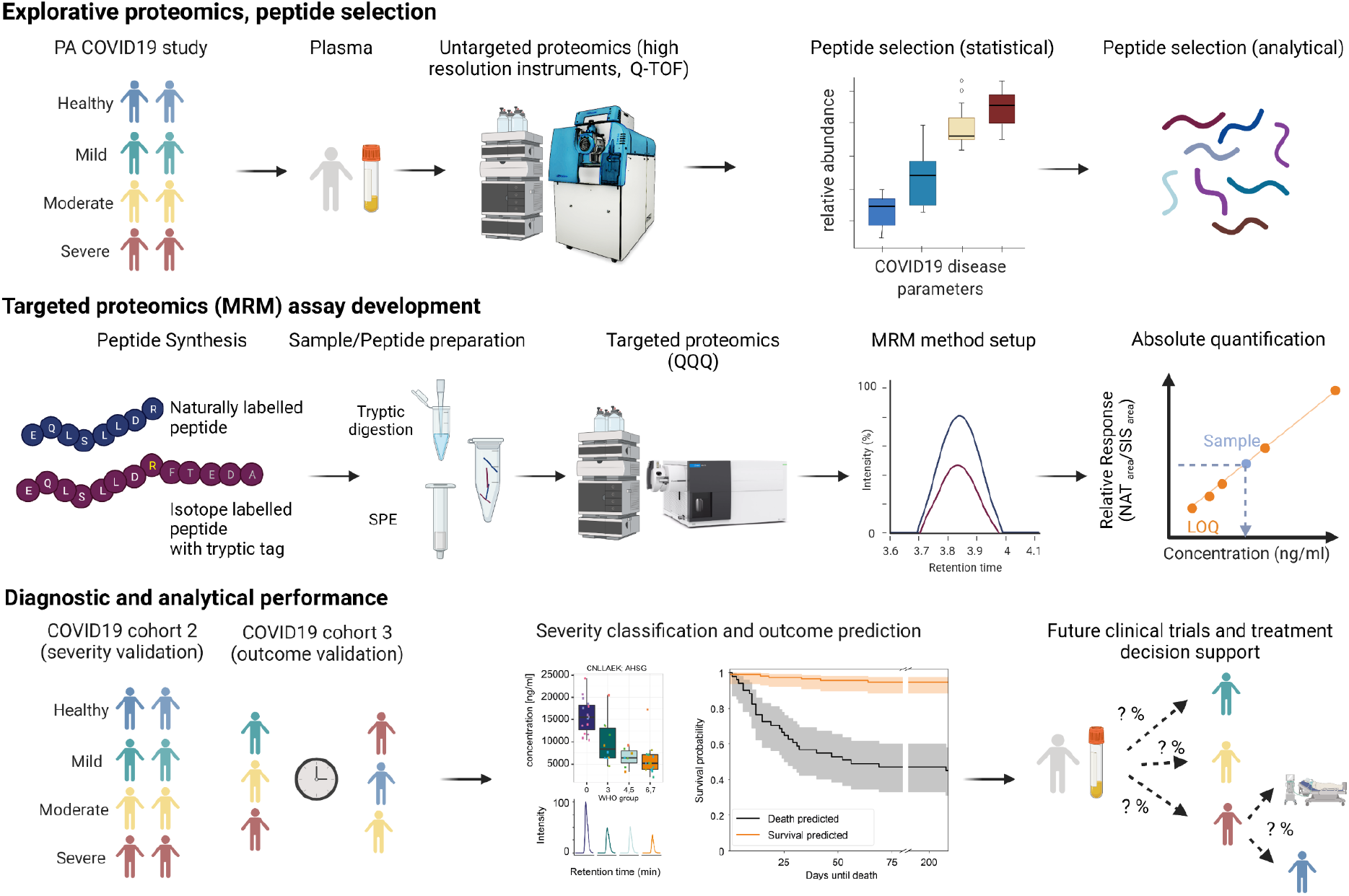
Schematic overview of the steps from peptide selection, to development and application of a COVID-19 biomarker panel for testing disease severity and predicting outcome. Top panel: Selection of 50 peptides derived from 30 plasma proteins as measured by discovery proteomics in a research setting [14,25,35] in the PA-COVID19 study cohort. Peptides were selected for a panel assay with the condition that they i) are prognostic about the remaining time in hospital in COVID-19 patients, ii) are differentially concentrated in the COVID-19 patient plasma according to the treatment escalation determined by the WHO ordinal scale, and iii) belong to biological processes that are causally implicated in the COVID-19 pathogenesis (including inflammatory response, complement cascade, metabolism, and the coagulation system). Middle panel: To generate an assay that is suitable for clinical and routine laboratories, we established a targeted LC-MRM assay to be used on conventional triple-quadrupole mass spectrometers, running routine-typical reversed phase chromatography, at a high flow-rate. The assay was optimized using synthetic peptides, and absolute quantification was based on corresponding stable isotope labelled internal standards (‘AQUA peptides [42]) with a short tryptic tag, to account for the sample preparation (tryptic digest) efficiency. Bottom panel: The assay was applied to two patient cohorts to evaluate the prognostic value of the assay for disease severity and outcome: a well balanced second (‘1st wave’) COVID-19 cohort [26], with samples being measured in two laboratories, and a larger longitudinal cohort (‘2nd wave’), with patients treated at the Charité Hospital, a national medical reference center.

**Figure 2.**
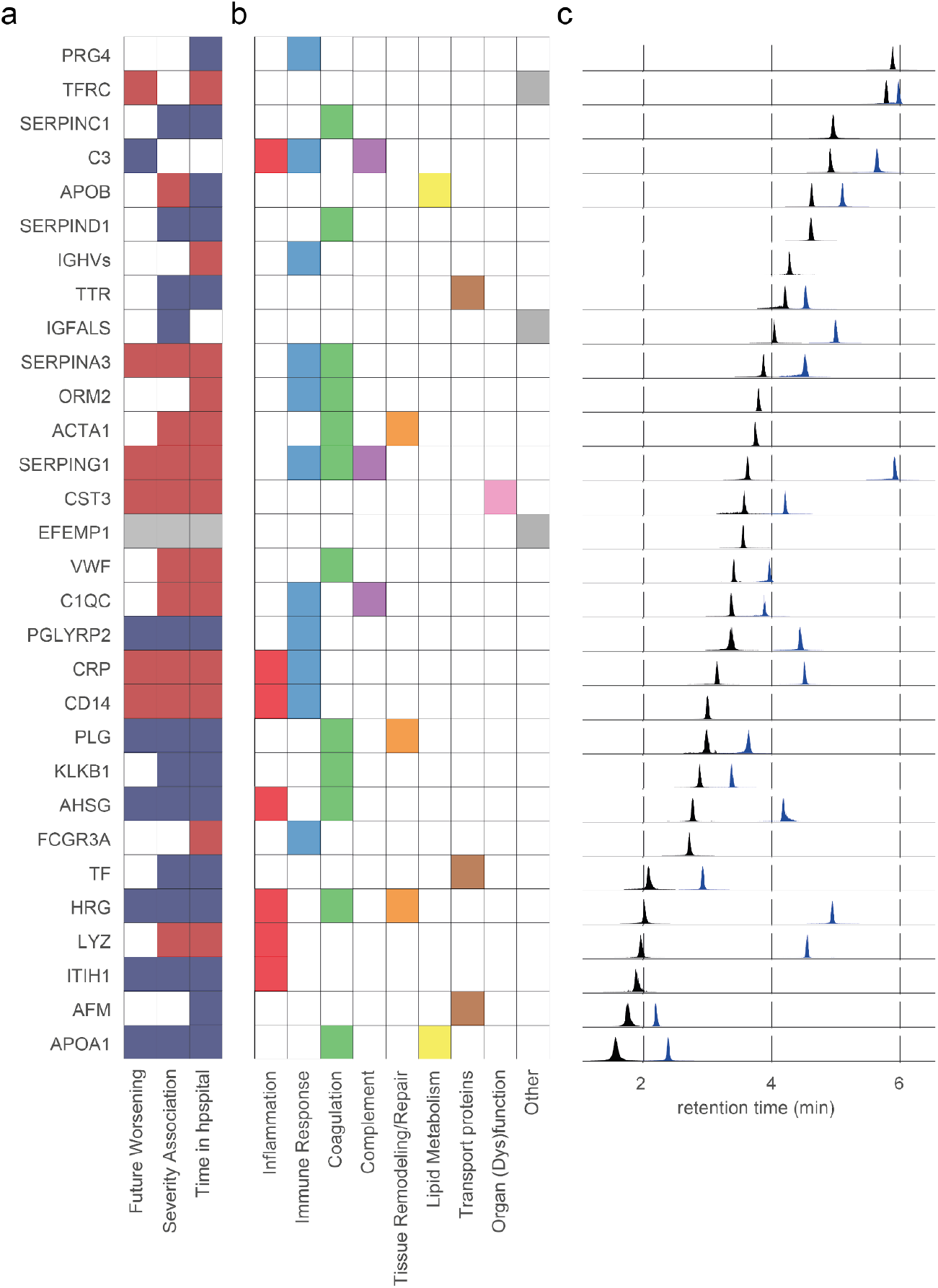
Selected protein biomarkers of the COVID-19 disease severity assay and their analysis by liquid chromatography, multiple reaction monitoring. **a)** Peptides selected for the panel assay are derived from proteins that are associated with COVID-19 disease parameters, including severity and progression. Colored tiles indicate significant associations, with red/blue highlighting that the respective protein is up- or down-regulated in COVID-19 infected individuals in discovery proteomic data [14]. **b)** Selected protein biomarkers and their associated COVID-19-pathology-related processes as curated from literature. **c)** Extracted ion chromatograms (EIC) highlighting the chromatographic spread of the applied MRM transitions selected as quantifiers for the indicated marker peptides. Each EIC was normalized to the maximum intensity of the respective peptide. Note that the majority of proteins are captured by two peptides (colored as blue, black for illustration purposes).

### Establishing an MRM based, targeted COVID-19 biomarker assay

We obtained synthetic reference standards for the 50 peptides, in both native and stable isotope labelled form. For each peptide 2 standards were synthesized: 1) with a natural isotope distribution (‘native’) and 2) with a C-terminal SIL amino acid (either [^13^C_6_,^15^N_2_]-lysine or [^13^C_6_,^15^N_4_]-arginine) to act as ISTDs, which contained a short tryptic tag to account for the digestion efficiency (Supplementary Table 1). The native peptides were employed to optimise liquid chromatography-mass spectrometry data acquisition method and quality of the Q1/Q3 (MRM) transitions. In order to establish the assay on a chromatographic system that could be run in a routine setting, we chose analytical flow-rate (800μl/min) reversed phase chromatography. We used a 1290 Infinity II (Agilent) UHPLC system, which was online-coupled to a triple quadrupole mass spectrometer (Agilent 6495C). Additionally, we set up a validation experiment in another laboratory and on another LC-MS/MS platform, for which an ExionLC AD UHPLC system was online coupled to another instrument (Sciex 7500, see below).

First, we set up the assay on the 6495C (Agilent) system. To select optimal MRM transitions for each peptide, we first predicted the transitions (consisting of several precursor ion charge states and respective product ions) using Skyline v21.1.0.146 [36]. We infused each native peptide solution into the LC-MS/MS systems and selected 1 precursor ion per peptide with the highest relative intensity and 5 most abundant product ions for collision energy optimisation as provided by Skyline. From these 5 product ions, we ultimately selected 2-5 experimentally optimised ion transitions per native peptide based on i) highest relative signal intensity, ii) optimal chromatographic peak shape and iii) absence of interfering signals. Product ions of <300m/z were excluded where possible to ensure specificity. Precursor and product ion-matched ISTD transitions were included. Lastly, all selected transitions were combined into one scheduled MRM method, which was subsequently analytically validated. The peptides were well distributed along a 8.6 minute linear gradient, and quantifiable with the chosen total runtime of 10 minutes (Fig. 2c). In the designed method, the most abundant transition for each peptide is used for quantification and 1-4 less abundant transitions are used as qualifiers (Supplementary Table 2).

### Technical validation of the peptide panel MRM assay

To evaluate the general applicability of the method for a clinical assay we tested performance of the selected peptides/transitions with respect to intra- and inter-batch repeatability, linearity, limits of quantification, accuracy, and potential matrix effects. First, we determined the intra- and inter-batch repeatability. We calculated the coefficient of variation (CV) for the triplicate of independently prepared calibration curves. These were constructed from serial dilutions of native peptide standards in BSA (40mg/ml), covering a concentration range ∼5×10^5^, and measured in technical pentuplicates (N=15) on the LC-MS/MS system. We used BSA as a surrogate matrix to test the analytical performance achieved on the standards in the absence of the endogenous plasma peptides [37] and achieved a median intra-batch CV of 2.6% and median inter-batch CV of 10.9% across low (LLOQ), medium ((LLOQ+ULOQ)/2) and high (ULOQ) concentration points (Table 1). The CV was determined from the response ratios, calculated by dividing native peptide peak area by internal standard peptide area. Additionally, we determined the limits of quantification (LOQ) as the highest and lowest concentration points on the linear calibration curve where the CV of the inter-batch repeatability was ≤20%. 37/50 peptides met or exceeded these LOQ criteria, as measured on the 6495C (Agilent) LC-MS/MS system. Since analytical validation requirements for clinical assays are purpose and context dependent, and are influenced by the magnitude of change of target analyte levels in control versus disease samples, we subsequently expanded the LOQ CV cutoff to ≤40%. This enabled us to determine LOQ of 10 additional peptides. Thus, we eventually determined limits of quantification and generated calibration curves for 47 peptides, with a median LLOQ of 1.6ng/ml. Obtained calibration curves also showed excellent linearity within the determined LOQs (R^2^ > 0.99) and typically spanned 3-4 orders of magnitude (Table 1).

As analytical technologies are sensitive to matrix effects [38], we evaluated parallelism in the surrogate BSA matrix compared to human plasma. We compared the slopes obtained from calibration samples measured in commercial human plasma samples (zenbio, Human Source Plasma, LOT#20CILP1034), with those measured in the surrogate matrix. 41/47 quantified peptide biomarkers that passed the above described validation showed no statistically significant matrix effects when comparing the slopes between matrices (P > 0.05) as expected for an assay using ISTDs [39,40]. For the 8 peptides (AADDTWEPFASGK, ADQVCINLR, ASDTAMYYCAR, ATEHLSTLSEK, DFALQNPSAVPR, EQLSLLDR, ESDTSYVSLK, IADAHLDR) that differed significantly a matrix factor (slope plasma/slope BSA x 100%) was calculated and reported (Table 1).

To test if peptide quantities from actual patient samples would be covered within the linear range of the calibration curves, we performed absolute quantification in plasma samples obtained from COVID-19 patients (pooled COVID-19 patient samples of different WHO treatment escalation grades; see Methods). Absolute quantification was performed by calculating the relative response ratio (peak area ratio of native peptide over its corresponding ISTD peptide) in pooled samples, followed by concentration interpolation from the above described BSA calibration curves. The peptide concentrations obtained from patient samples were covered within the determined linear range of the assay and were measured with a median accuracy of 97.3% across low (LLOQ), medium ((LLOQ+ULOQ)/2) and high (ULOQ) concentration points (Table 1).

### The COVID-19 MRM panel assay reports disease severity in an early pandemic cohort

Next, we assessed how the absolute concentration of the quantified biomarkers changed as a function of the COVID-19 treatment escalation level, a proxy for disease severity, as expressed by the 7-point WHO ordinal scale of clinical improvement [33]. To establish this relationship, we applied the panel assay on plasma samples obtained from a deeply characterized, early pandemic COVID-19 cohort, hospitalized during the first wave of the pandemic between 1st and 26th of March 2020 (‘Cohort 2’, n=45) [26]. This cohort was ideally suited for this validation step, as it was well balanced from patients with mild to severe COVID-19 and included healthy controls [26,32]. Furthermore, the cohort was sampled as citrate plasma, in difference to the exploratory cohorts in our previous studies that we used for identifying the biomarkers which were sampled as EDTA plasma. This test was hence also indicative if our choice of biomarkers would allow stable conclusions across alternative sample matrices.

For robust and reliable proteomic sample preparation, we employed a recently presented procedure that enables tryptic digest and solid phase extraction in a semi-automated way [25]. 40/50 peptides could be reliably detected and quantified in the patient citrate plasma (Supplementary Fig. 1). The concentration of 32 peptides changed with the severity of COVID-19 according to treatment escalation: i.e. from uninfected (WHO 0) to mildly (WHO 3), moderately (WHO 4, 5) and severely (WHO 6, 7) affected COVID-19 patients (Fig. 3a, Supplementary Fig. 1, P < 0.05). Most of the chosen markers change in abundance between healthy and COVID-19 infected individuals, and further follow their respective trend with increasing treatment escalation level, such as peptides derived from the acute phase proteins CRP and AHSG, or the innate immune response protein PGLYRP2 (Fig. 3a, Fig. 3b). Some of the peptides give a signal during specific disease state transitions, in that they differ between an infected and uninfected individual, such as peptides from the complement related protein SERPING1 or the iron-binding protein TF (Fig. 3a, Fig. 3b), or change the most during the most severe treatment escalations of COVID-19, such as the kidney- and inflammation-marker CST3 (Fig. 3a, Fig. 3b). Indeed, despite the comparatively small scale of this cohort, the obtained peptide-abundance profiles did distinguish the COVID-19 patients according to the necessary treatment level (WHO score), indicating the panel captures disease severity (Fig 3c). In summary, we successfully quantified 40 peptides in COVID-19 patient citrate plasma samples by high-flow LC-MRM. 32 of these peptides were differentially concentrated depending on disease severity in a COVID-19 inpatient cohort, and did classify the patients according to their treatment escalation.

**Figure 3.**
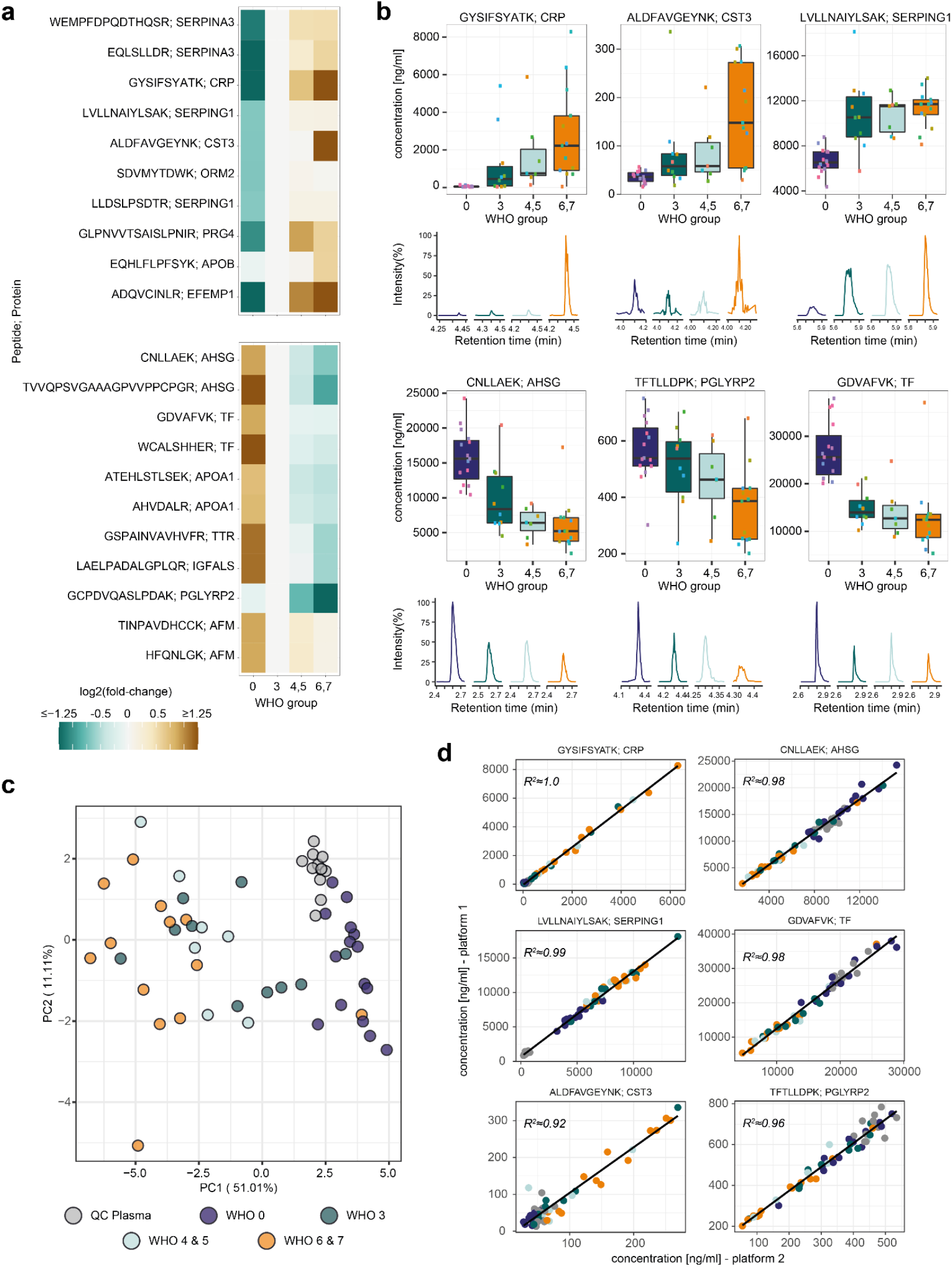
The protein biomarker assay reports disease severity on a balanced COVID-19 cohort, and produces reproducible results in two laboratories using different LC-MS/MS platforms. **a)** The established assay was applied to citrate plasma samples, collected for a balanced COVID-19 cohort studied during the first wave of the pandemic [25,26,32] (‘cohort 2’) consisting of healthy volunteers (n=15, WHO grade 0), COVID-19 affected individuals requiring hospitalisation but no oxygen therapy (n=10 (WHO 3), COVID-19 affected individuals requiring hospitalisation and non-invasive oxygen therapy (n=4, WHO 4; n=3 WHO5), and severely affected hospitalised individuals requiring mechanical ventilation (n=3 (WHO 6), n=10 (WHO7) as well as QC plasma samples (n=12). Peptides with a significant concentration change (up- (top panel) and down-regulated (bottom panel)) distinguish healthy from infected individuals, as well as mild from severe forms of the disease. Heatmap displays the log2 fold-change of the indicated peptide to its median concentration in patients with a severity score of WHO3. To facilitate visualisation, peptides with log2 fold-changes <-1.25 or >1.25 are indicated by the same respective color. Peptides are ordered based on significance, with most significant peptides of each respective panel on top. Only peptides with P < 0.05 based on Kendall’s Tau trend estimator corrected for multiple testing are shown. For additional information, see Supplementary Fig. 1. **b)** Visualisation of the response to COVID-19 based on selected peptide biomarkers indicating different COVID-19 severity trends (changing with severity expressed according to the WHO ordinal scale (left, middle panel), and differentiating healthy from COVID-19 infected individuals (right panel)). Boxplots display the absolute concentration of selected peptides in patients in different severity groups as explained in (a). The extracted ion chromatograms (EIC) display the response of representative samples of individuals classified according to the treatment escalation WHO=0, WHO=3, WHO=5 and WHO=7. **c)** Unsupervised clustering as visualised with a principal component analysis (PCA) based on the absolute concentration of 39 quantified peptides clusters COVID-19 patients by severity. The peptide ADQVCINLR contained missing values and was omitted. **d)** Analytical reproducibility of the assay in two laboratories, running two different LC-MS/MS platforms, with independently optimised MRM transitions. Shown are linear correlations between peptide quantities. Selected peptides and color code like in (c).

### Analytical cross-platform and cross-laboratory validation

To evaluate transferability of the assay, samples from the above described cohort (cohort 2) were measured on both the 6495C (Agilent, Supplementary Table 2) and the 7500 (SCIEX, Supplementary Table 3) LC-MS/MS platforms, in different laboratories. For 33/40 selected peptides, we obtained an excellent cross-laboratory/cross-instrument correlation between the concentration measured in respective COVID-19 patient samples (Fig. 3d, Supplementary Fig. 2). One peptide (ESDTSYVSLK) suffered from one outlier, but otherwise had a good correlation on both platforms (Supplementary Fig. 2). The remaining (six) peptides were close to the detection limit in the citrate plasma sample matrix, which likely explains the higher variance and limited correlation (R^2^ < 0.6) between both platforms. Further, on a subset of peptides we observed an excellent correlation but different absolute values, which points to differences in the calibration. Despite this, principal component analysis revealed near identical sample group clustering on both platforms (data not shown). Overall, as we detected and quantified the majority of peptides on both platforms with high precision, the peptides that differentiate between different stages of COVID-19 can be quantified on both platforms, demonstrating the general applicability of the assay independent of the employed analytical instrumentation.

### Severity stratification and prediction of disease progression in a longitudinal COVID-19 cohort

We next applied the panel assay on a larger, longitudinal collection of samples obtained during the second wave of the pandemic in Germany (04 April to 19 November 2020). This cohort (cohort 3, 165 patients included) was selected i) because the large number of samples (n=552 samples from 165 patients) add information about the technical stability of the assay and increase the statistical power to evaluate the results, and ii) the longitudinal nature of the study allowed us to assess a potential prognostic value of the MRM panel assay.

Reassuringly, despite the large number of proteome samples acquired split over three batches and measured over 10 days, and the different matrix (EDTA plasma), technical variation was low (Fig 4a). The abundance of peptides which we had found characteristic for disease severity in cohort 2 (Fig 2b) clearly differed between different WHO grades, whereas the QC measurements were highly reproducible (Fig. 4b).

**Figure 4.**
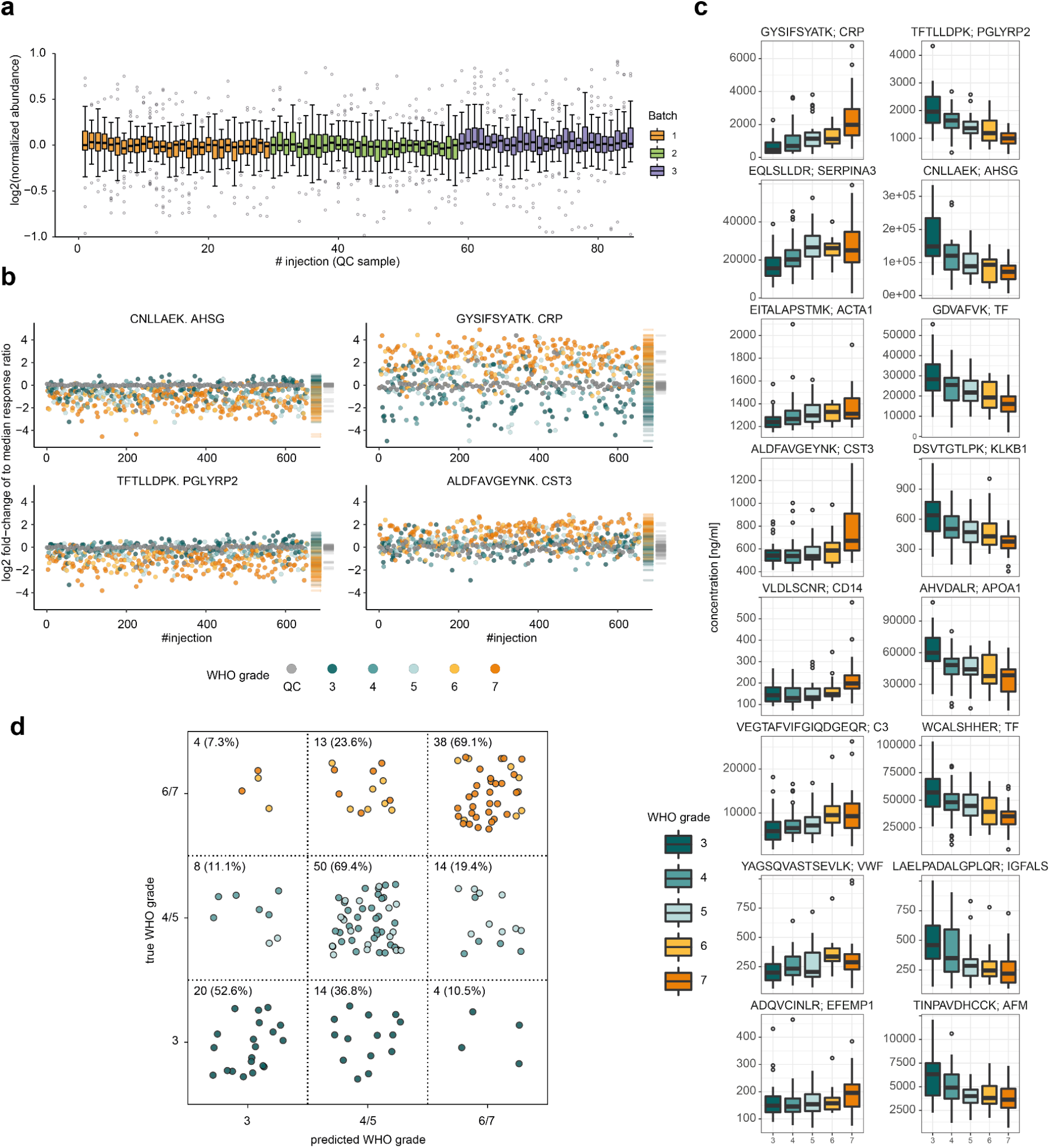
Diagnostic and analytical performance of the multi-protein panel assay on a COVID-19 inpatient cohort treated during the 2nd wave of the pandemic. **a)** Quantitative performance (signal stability), during the measurement of 552 plasma proteome samples of patient cohort 3 evaluated based on n=85 QC samples (colored in grey, pool of COVID-19 samples as described in [25]) injected throughout the acquisition. Shown are the log2 fold-change of the response ratios (peak area of naturally- over isotope-labelled peptides for each of the 48 quantified peptides) normalised to the median of the QC samples for the respective peptide. **b)** Peptide log2 response-ratios of two selected down- (AHSG, PGLYRP2) and two up-regulated (CRP, CST3) proteins for all samples acquired for the cohort described in (a). QC samples are shown in grey, all other samples are colored according to the corresponding COVID-19 WHO treatment escalation score; rug plots on the right side of each peptide indicate the respective distributions. **c)** Illustration of the severity stratification ability; the 8 most significantly upregulated (left panel) and down-regulated (right panel) peptide quantities that indicate COVID-19 disease severity, expressed as the treatment level according to the WHO scale. Only one peptide per protein was selected; one outlier sample in peptide EITALAPSMK was removed for visualisation purposes only. The quantities of all quantified peptides are illustrated in Supplementary Fig. 3. **d)** Confusion-matrix-like representation of the outcome of a multi-class classification model (SVM-based) trained to differentiate three WHO severity groups: grade 3, grades 4/5, and grades 6/7. Predictions were done on withheld samples that were not used for training the models (accuracy = 0.655, balanced accuracy = 0.637). The percentage denotes how many samples within each WHO severity group are assigned to each square. The positions of the points within each square were chosen randomly. Color scheme according to (c).

To evaluate the ability of the MRM panel to differentiate COVID-19 severity, we first analyzed the earliest sample obtained for each patient of this cohort. Not only is the earliest sample during disease progression of highest practical value, it is also the furthest apart from outcome, and hence best to validate if there is a prognostic value in measuring the biomarker panel. In EDTA plasma, the assay quantified 47/50 peptides. The majority of these (33 peptides) had a significantly different trend between patients according to their treatment escalation level (WHO3 to WHO7), with 11 peptides significantly increasing, and 22 decreasing in concentration (Fig. 4c, Supplementary Fig. 3). Further, we again observed peptide abundance profiles indicative of specific disease state transitions related to the treatment escalation; for instance, the abundance of peptides corresponding to the proteins CRP, CST3, or CD14 were especially increased in very severe forms of the disease (WHO grade 7) (Fig. 4c). This analysis hence confirmed that the MRM panel reports disease severity in COVID-19, in two matrices (citrate and EDTA plasma) and two cohorts, sampled at a different stage of the pandemic.

Next, we tested if the necessary treatment level could be predicted from the first time point sample available. We constructed a support vector machine (SVM) trained to differentiate between three different treatment groups on the basis of the severity markers: WHO grade 3 (mild COVID-19, hospitalised, but no supplemental oxygen necessary), WHO grade 4/5 (moderate COVID-19, hospitalised, supplemental low- or high flow oxygen necessary) and WHO grade 6/7 (severe COVID-19, hospitalised, intensive care and invasive mechanical ventilation necessary). The data was split in a training and a validation set in a cross-validated manner. The model accurately predicted the WHO grades in the validation set from the peptide biomarker data (Fig. 4d). This performance strengthens the observed significant differentiation of WHO grades based on single peptides and shows that combinations of these peptides are capable of predicting disease severity groups.

An important clinical need for a COVID-19 panel assay would be to be prognostic about outcomes, as established risk assessments such as the SOFA, APACHE II and CCI can have a moderate performance on severe COVID-19 patients [35]. As disease severity correlates with outcome (i.e. a patient that is severely ill has a much higher chance to die from COVID-19 than a mildly affected patient) we hence asked if the selected biomarker panel would be prognostic. An SVM was trained on data obtained from the earliest sample, in a cross-validated manner, to differentiate patients who later survived COVID-19, from patients with a fatal outcome (n=165, of which 131 survived (controls) and 34 died (cases)) (Fig. 5). The trained outcome-predictor was able to correctly classify 82.4 % of the patients (sensitivity = 0.824, (28/34) specificity = 0.824 (108/131), AUROC = 0.872) that were withheld while training the model (Fig. 5a, b). To exclude that the predicting capabilities are limited to the method, another predictor (extra-trees) using the same setup was evaluated (Supplementary Fig. 4). Reassuringly, this predictor (sensitivity = 0.735, specificity = 0.817, AUROC = 0.844), shows a comparable performance. To evaluate how well the SVM-predictor performs compared to the clinically established scores, we determined the SOFA, APACHE II, CCI scores, all of which are in clinical use. SOFA and the APACHE II, which are directly linked to the severity of the patient, performed best among the three conventional scores tested. Nonetheless, the MRM biomarker assay significantly outperformed all three clinical scores (AUC of the ROC-curves) (Fig. 5a).

**Figure 5.**
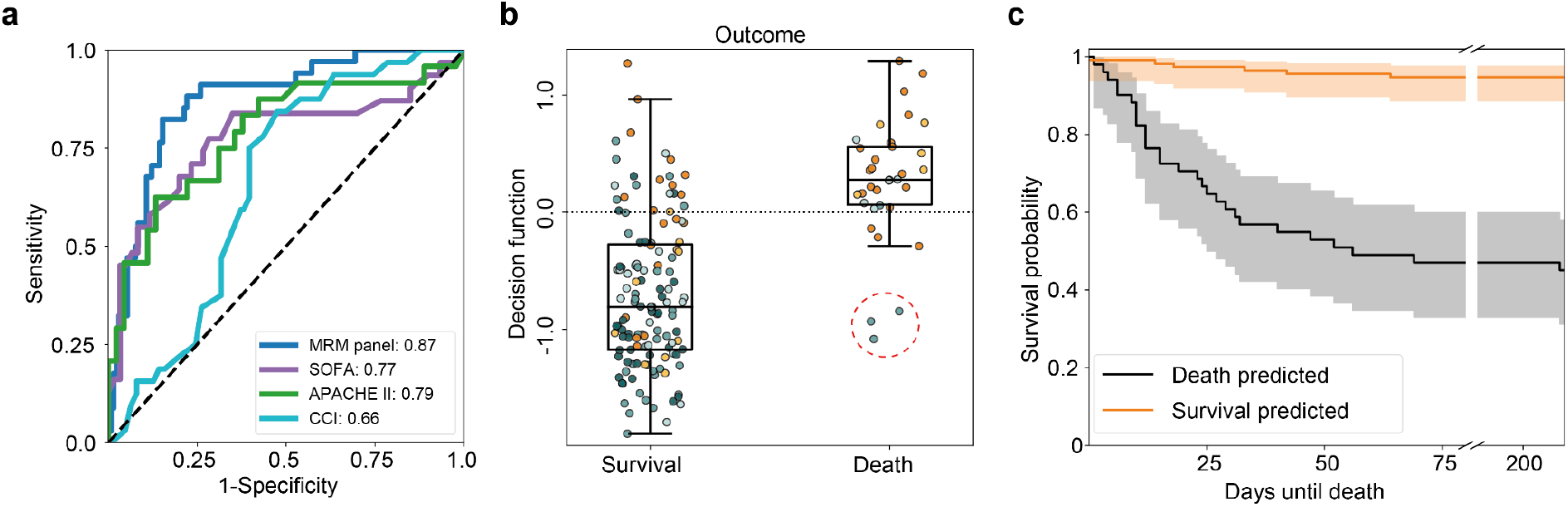
COVID-19 outcome prognosis using a routine applicable disease severity LC-MRM assay. **a**) Receiver operating characteristic (ROC) curve for the prediction of survival and non-survival, from a single plasma sample (first time point measured for every patient) using an SVM-classifier. The blue curve denotes the model trained and benchmarked on measured proteomic data. The other curves denote models based on single severity scores (Sepsis-related Organ Failure Assessment score (SOFA, purple), Acute Physiology And Chronic Health Evaluation (APACHE II, green) and Charlsson Comorbidity Index (CCI, cyan)). **b)** Boxplot of the decision function of the SVM for every patient sorted according to the outcome and colored with respect to the WHO grade at the day the sample was taken. Color scheme according to Fig. 4c. The red dashed circle indicates three patients with a high chance of predicted survival who died. These patients all were WHO4 patients with ‘do not intubate (DNI)’ orders in place due to other medical conditions. Therapy was therefore not escalated to invasive ventilation. The MRM assay classified those patients as milder COVID-19 cases, compared to other non-survivors. **c)** Kaplan-Meier estimate of the survival function for survival predicted cases (orange) or non-survival predicted cases (black) with confidence interval (alpha=0.05). All predictions were done on withheld samples that were not used for training the models.

## Discussion

Novel infectious diseases such as COVID-19 that lack immediate treatment options can quickly challenge health systems on a global scale. Accordingly, there still exists a substantial and unmet clinical need for assays that capture and monitor the individual response of patients. Such personalized tests can support clinical decision making and guide development of novel treatments. For this reason, a range of clinical scoring systems have been conceived and tested for their application in COVID-19 [20,21]. Several investigations have highlighted the prognostic value of plasma proteomes in COVID-19, and indicated that proteomics could outperform existing tests [10,11,18,22–24]. However, discovery proteomic technologies are difficult to establish in clinical routine due to technical reasons, throughput and cost. The most efficient way to translate a proteomics result is hence through the selection and validation of biomarker panels that can be measured on established and certified routine laboratory equipment.

The here presented test relies on a multi-protein biomarker panel for severity stratification in COVID-19. The target peptides were selected based on discovery proteomic data recorded with SWATH-MS [14,25,26], and used in the development and validation of a clinically translatable and scalable LC-MRM based assay. We choose targeted mass spectrometry in the form of MRM, which in other research and clinical areas (such as newborn screening, toxicology) is established as a paramount methodology for the quantification of multiplexed or multiparametric marker panels in biofluids [41]. LC-MRM is an ideal technology in this application as it i) provides excellent sensitivity and specificity ii) has the possibility to easily include internal standards that give the assay precision and enable control over potential matrix effects [39,40], iii) facilitates absolute quantification, enabling cross-platform transferability [42,43] and iv) covers a large dynamic range (3-4 orders of magnitude of linear range in the presented assay) [44]. This in turn enables the comparison of biomarkers with large abundance differences within one run, thus facilitating multiplexing of many biomarkers in parallel, even without the need for matrix depletion [45]. Given the simplicity, flexibility, and multiplexability of MRM-based LC-MS/MS assays, the initial panel of peptides can be large, increasing the chance of finding a reliable set of reproducible biomarkers based on hierarchical filtering and selection of the most suitable candidates during establishment of the targeted assay [46,47]. Finally, although LC-MRM assays require analytical proficiency and are labor-intensive to set up, they are highly cost-effective to run and can be adapted directly by routine clinical laboratory scientists to meet a change in needs. In our study, we overcame a common limitation of proteomic assays for their routine use - their dependency on low flow rate-chromatography [48–50]. Exploiting the high sensitivity of contemporary triple-quadrupole mass spectrometers, we demonstrate the accurate quantification of the peptide panel using analytical flow rate chromatography, which not only is robust and fast, but also routinely used in clinical laboratories for small molecule analysis, greatly simplifying the application of our assay in the clinical routine.

The developed biomarker panel includes 50 peptides derived from 30 plasma proteins. The proteins are related to biological processes which have been shown to be important for the COVID-19 host response and pathophysiology, like the innate immune response, the coagulation system or the complement cascade [14,25,26,35]. The assay is hence monitoring processes that are closely linked to disease progression and the exceptionally diverse clinical presentation of patients with SARS-CoV-2 infection. In this study, we established the assay on two routine-laboratory-compatible LC-MRM platforms, and performed analytical validation of the key technical aspects of the assay. We demonstrate excellent sensitivity, accuracy, precision, as well as reproducibility across two different targeted mass spectrometry platforms. Of note, reagents for sample preparation and LC-MRM method setup are also commercially available in standardised kit formats. As such, the assay could be easily translated into a multiplexed, high-throughput, standardised and scalable clinical assay that captures the hallmarks of COVID-19 pathophysiology.

We confirm in two temporally separated COVID-19 patient cohorts that the panel assay accurately captures disease severity of SARS-CoV-2 infected individuals and discriminates the necessary treatment levels. As disease severity correlates with outcome, we also tested the prognostic value of the panel and found that it outperformed three clinically established risk assessment metrics, CCI, SOFA and APACHE II, in predicting the survival of COVID-19 inpatients. Thus, the panel assay parameters could be used to assess the current state of the patient, help to monitor novel treatments, or stratify patients based on their responsiveness to novel therapeutic interventions. Furthermore, the assay can be employed to predict the future course of COVID-19, as exemplified by the prediction of disease outcome weeks into the future. It could therefore potentially help to guide clinical decision making, and support resource allocation, particularly in critical situations of the pandemic with overstrained healthcare capacities. While the value of quantitative proteomic measurements for disease stratification and prediction could be validated in different cohorts, not all eventualities of a complex disease such as COVID-19 are necessarily covered. To this end, additional data, and ideally prospective studies will be of high value, especially to refine predictive models, and to assess the performance of the assay in various populations.

The biomarker panel was primarily selected for measuring disease severity in all inpatient groups, ranging from a mild to a severe form of the disease. In a parallel study, we have used discovery proteomics, and found that proteomics can also be prognostic for outcome in patients with similar disease severity, e.g. among severely affected patients, that are difficult to distinguish by clinical parameters [35]. This means that the prognosis of survival using targeted proteomics could be improved beyond what was shown in this study for ‘within-severity-group’ prognosis, if biomarker panels specifically for stratification within the respective COVID-19 severity group would be selected. Indeed, particularly within the group of severely affected individuals, some patients got predicted wrongly, i.e. survived despite being predicted as non-survivors, or vice versa (Fig. 5b).

We assessed on a patient-by-patient basis whether there are medical reasons that could explain wrong predictions. Plotting outcome with respect to the time until death (Kaplan-Meier survival analysis, Fig. 5c) denotes no clear tendency for the correct and false predictions. However, we noted that the three samples with the smallest decision function across wrongly predicted patients with fatal outcomes belonged to WHO 4 patients that had DNI (‘do not intubate’) orders in place (denoted with a red circle in Fig. 5b). It is hence plausible that the assay correctly identified a milder form of COVID-19 in these three individuals; i.e. that without a strong comorbidity or a DNI order in place, these might have had a good chance to survive COVID-19.

COVID-19 will remain a central public health issue for the foreseeable future as new variants of concern with capacity to evade vaccine induced immunity continue to emerge [9]. The presented peptide panel utilises only 5μl of plasma enabling potential finger-prick microsampling, which would in turn enable even remote patient monitoring at home. Further, the underpinning targeted proteomics platform supports rapid (45-60 days) iteration of the panel composition in case additional prognostic biomarkers are discovered, for instance in additional patient cohorts. Taken together, this peptide panel and the underlying analytical platform, holds potential to support a broader, continuous pandemic response in addition to its utility in hospitalised patient cohorts which we demonstrate in the present study.

## Data Availability

All data produced in the present study are available upon reasonable request to the authors.

## Acknowledgements

This research was funded in part by the European Research Council (ERC) under grant agreement ERC-SyG-2020 951475 (to M.R), the Wellcome Trust (IA 200829/Z/16/Z to M.R.). The work was further supported by the Ministry of Education and Research (BMBF), as part of the National Research Node ‘Mass spectrometry in Systems Medicine (MSCoresys), under grant agreements 031L0220 and 161L0221. J.H. was supported by a Swiss National Science Foundation (SNF) Postdoc Mobility fellowship (project number 191052). This study was further supported by the German Federal Ministry of Education and Research (NaFoUniMedCOVID-19 – NUM-NAPKON, FKZ: 01KX2021). The study was co-funded by the UK’s innovation agency, Innovate UK, under project numbers 75594 and 56328. Fig. 1 was made using BioRender (Biorender.com). The Pa-COVID-19 study group is acknowledged for study logistics and collection of biosamples and clinical data.

## Methods

### Patient cohorts

Patient samples were collected as described previously [14,25,26,35] as part of a prospective observational cohort study Pa-COVID-19 at Charité – Universitätsmedizin Berlin. The study protocol has been described in detail before [32]. The study is registered in the German and the WHO international registry for clinical studies (DRKS00021688). The cohorts are summarized in Supplementary Table 4.

### Reagents and peptide standards

Reference peptide standards were custom synthesised where native peptides were obtained at ≥95% purity and SIL internal standard peptides - at ≥70% purity. Internal standards contained 4-6 amino acid tryptic tags mimicking the sequence in a corresponding human plasma protein and were labelled on C-terminal lysine (K) or arginine (R) amino acids with stable isotopes (K(U-^13^C_6_,^15^N_2_) or R(U-^13^C_6_,^15^N_4_)). Water was from Merck (LiChrosolv LC-MS grade; Cat# 115333), acetonitrile was from Biosolve (LC-MS grade; Cat# 012078), trypsin (Sequence grade; Cat# V511X) was from Promega, 1,4-Dithiothreitol (DTT; Cat# 6908.2) from Carl-Roth, iodoacetamide (IAA; Bioultra; Cat# I1149) and urea (puriss. P.a., reag. Ph. Eur.; Cat# 33247) were from Sigma-Aldrich, ammonium bicarbonate (Eluent additive for LC-MS; Cat# 40867) and Dimethyl sulfoxide (DMSO; Cat# 41648) were from Fluka, formic acid (LC-MS Grade; Eluent additive for LC-MS; Cat# 85178) was from Thermo Scientific™, bovine serum albumin (BSA; Albumin Bovine Fraction V, Very Low Endotoxin, Fatty Acid-free; Cat# 47299) was from Serva.

All peptide stock solutions were prepared at 1 mg/ml concentration in 50:50 ddH_2_O: acetonitrile mix, except for STDYGIFQINSR and VEGTAFVIFGIQDGEQR where 200μl of DMSO were added to solubilise the peptides at 5mg/ml which were then aliquoted and diluted to 1mg/ml with 50:50 ddH_2_O: acetonitrile mix. Internal standard mix was prepared by pooling 20μl of each stable isotope-labeled peptide, evaporating 200μl of this mix to dryness and reconstituting in a denaturation buffer to the final concentration of 1.4 μg/ml for each peptide. Cassetted calibration curves were prepared by serial dilution of pooled native reference peptide standards as described in *Analytical method validation*. After serial dilution, these samples were treated identically to respective clinical samples.

### Sample preparation

Samples were prepared as described previously (Messner et al., 2020) with minor modifications. Briefly, clinical samples and calibration lines in Cohort 2 were prepared as follows: 5μl of citrate plasma sample were added to 55μl of denaturation buffer, composed of 50μl 8M Urea, 100mM ammonium bicarbonate, 5μl 50mM dithiothreitol (DTT) and peptide internal standard mix. The samples were incubated for 1h at room temperature before addition of 5μl of 100mM iodoacetamide (IAA). After a 30min incubation at room temperature the samples were diluted with 340μl of 100mM ammonium bicarbonate and digested overnight with 23μl of 0.1μg/μl trypsin at 37°C. The digestion was quenched by adding 50μl of 10% v/v formic acid. The resulting tryptic peptides were purified on a 96-well C18-based solid phase extraction (SPE) plate (BioPureSPE Macro 96-well, 100mg PROTO C18, The Nest Group). The purified samples were resuspended in 120μl of 0.1% formic acid and 20μl or 0.2μl were injected into 2 LC-MS/MS platforms (6495C and 7500 respectively).

Samples in cohort 3 were prepared as described above, with one modification. EDTA plasma was used instead of citrate plasma, and internal standards were digested separately and added to pre-digested clinical and calibration line samples before their injection into the LC-MS/MS system. Quality control (QC) samples consisted of pooled commercial control and COVID-19 human plasma samples (as described in a previous publication [25]), and were prepared alongside clinical and calibration curve samples in each cohort.

The COVID-19 sample pools used for the analytical validation were generated by pooling 5μl of patient plasma from cohort 3 according to their WHO treatment severity score. Only samples of patients that had not received dexamethasone were used.

### Liquid chromatography - tandem mass spectrometry

Tryptic peptides were quantified on 2 liquid chromatography - triple quadrupole mass spectrometry (LC-MS/MS) platforms - 6495C (Agilent) and 7500 (SCIEX).

#### 6495C (Agilent) LC-MS/MS method

All clinical samples were analysed on the Agilent 6495C mass spectrometer, coupled to an Agilent 1290 Infinity II UHPLC system. Prior to MS analysis, samples were chromatographically separated on an Agilent InfinityLab Poroshell 120 EC-C18 1.9μm, 2.1×50 mm column heated to 45°C and with a flow rate of 800 μl/min. Linear gradients employed were as follows (time, % of mobile phase B): 0 min, 3%; 1min, 3%; 7.5min, 35%; 8min 98%; 8.5min, 98%; 8.6min, 3%; 10min, 3% where mobile phase A & B are 0.1% formic acid in water and 0.1% formic acid in acetonitrile respectively.

The 6495C mass spectrometer was controlled by Agilent’s MassHunter Workstation software (LC-MS/MS Data Acquisition for 6400 series Triple Quadrupole, Version 10.1) and was operated in positive electrospray ionisation mode with the following parameters: 3500 V capillary voltage (positive), 0 V nozzle voltage (positive), 12 L/min sheath gas flow at a temperature of 280°C, 17 L/min gas flow at a temperature of 170°C, 40 psi nebulizer pressure, 166 V fragmentor voltage, 5 V cell accelerator potential. Samples were analysed in dynamic MRM mode with both quadrupoles operated in unit resolution. All other MRM parameters, including monitored transitions and scheduling are provided in the Supplementary Table 2.

#### 7500 (SCIEX) LC-MS/MS method

Samples from cohort 2 were analysed on a SCIEX 7500 mass spectrometer coupled to an ExionLC AD UHPLC system (SCIEX, UK) in addition to the analysis on the Agilent platform. Prior to MS analysis, samples were chromatographically separated on a Phenomenex Luna Omega Polar 3 μm, 100 × 2.1 mm column heated to 40 °C and with a flow rate of 500 μl/min. Linear gradients employed were as follows (time, % of mobile phase B): 0 min, 3%; 0.1min, 3%; 7.5min, 30%;8min 95%; 8.5min, 95%; 8.6min, 3%; 10min, 3% where mobile phase A & B are 0.1% formic acid in water and 0.1% formic acid in acetonitrile respectively.

The 7500 triple quadrupole mass spectrometer was operated in positive electrospray ionisation mode with the following ion source parameters: 1750 V Ionspray voltage, 40 psi curtain gas, 40 psi Ion source gas 1, 70 psi ion source gas 2 and 500 °C temperature. Samples were analysed in Scheduled MRM mode with both quadrupoles operated in unit resolution. All other MRM parameters, including monitored transitions and scheduling are provided in the Supplementary Table 3.

### Mass spectrometry data processing

Mass spectrometry data processing was performed with vendor-specific software: Agilent MassHunter Quantitative Analysis, v10.1 and SCIEX OS Software v2.0.1. Peak selection and integration were manually assessed before exporting the peak area values to .csv for further analysis. Peptide absolute concentration was determined from calibration curves, constructed with native and SIL synthetic reference standards. Of note, SIL internal standards for 5 corresponding native peptides could not be detected on the 6495C system. To quantify these native peptides, we used other, closely eluting SIL internal standards in the assay: AADDTWEPFASGK(U-13C6,15N2 was used for ASDTAMYYCAR, GYSIFSYATK(U-13C6,15N2) for GSPAINVAVHVFR and WEMPFDPQDTHQSR, ANRPFLVFIR(U-13C6,15N4) for LAELPADALGPLQR and VSASPLLYTLIEK(U-13C6,15N2) for VEGTAFVIFGIQDGEQR. In addition, due to low signal intensity of pre-assigned quantifier transitions (transitions with matched precursor and product ions across native and SIL peptides), we chose other transitions with higher signal intensity for 4 SIL peptides, even if they did not match the fragmentation pattern of their respective native peptides. The transitions used for quantification on both 6495C and 7500 LC-MS/MS platforms are shown in Supplementary Tables 2 and 3 respectively.

Linear regression analysis of each calibration curve was performed using custom R code or SCIEXOS (with 1/x weighting). The respective peptide concentration in patient samples is expressed in ng/ml.

### Analytical method validation

Method analytical validation was performed based on FDA Bioanalytical Method Validation criteria [51] where sensitivity, specificity, intra, inter-repeatability, accuracy and matrix effects have been assessed. A total of 5 independent calibration curves were prepared by serial dilution of native reference peptide standards in assay buffer (1), surrogate matrix (3) and pooled human plasma (1) across the final sample peptide concentration range of 0-1.63μg/ml. Surrogate matrix (40 mg/ml BSA) calibration curves were prepared and analysed across 3 separate batches and all calibration curve samples were analysed in quintuplets. Linear 1/x weighted calibration curves were obtained for all calibration curves in order to check the linearity of the response. Lower limit of quantification (LLOQ) was defined as the lowest calibration sample on the linear curve with a CV ≤ 20%. Ten peptides where low endogenous concentrations were observed in a pooled clinical subset of samples, LLOQ criteria were expanded to CV ≤ 40% to prevent missing values where these peptides that were highly differentially expressed between COVID-19 severity groups were successfully quantified. Upper limit of quantification (ULOQ) was defined as the highest calibration sample on the linear curve with a CV ≤ 20%.

Accuracy was assessed by treating 1 of the 5 replicates in each calibration curve in the surrogate matrix as pseudo-unknown samples, quantifying with the curve generated from the remaining 4 replicates and calculating a median accuracy of all replicates. Matrix effects were measured by comparing the slopes of calibration curve samples prepared in a BSA matrix and pooled human plasma. Here an Extra Sum of Square F test was used for statistical comparison with a p-value < 0.05 indicating potential matrix effects.

### Data analysis and visualisation, statistics

Significance testing of the trend between absolute peptide concentrations and the ordinal classification as provided by the WHO treatment escalation scale (levels as indicated) was performed using Kendall’s tau (KT) statistics as implemented in the “EnvStats v2.4.0” R package “kendallTrendTest” function. For cohort 2 the KT statistics was calculated as the trend of peptide quantities against the following WHO groups: 0, 3, 4, 5, 6, 7; for cohort 3, peptide quantities were tested against the following WHO groups: 3, 4, 5, 6, 7. A full summary of statistical test results is provided in Supplementary Table 5. Multiple testing correction was performed by controlling for false discovery rate using the Benjamini-Hochberg procedure [52] as provided by the R package “stats v4.1.0” - “p.adjust” function. Principal component analysis was performed using the R function “prcomp” from the “stats 4.1.0” package and visualized using “ggplot2 v3.3.5”.

### Prediction of WHO grade and disease outcome

For the prediction of the current WHO grade and for the outcome prediction a Support Vector Machine was used as implemented in *scikit-learn 0*.*23*.*2* (*sklearn*.*svm*.*SVC*) [53] using default parameters (rbf-kernel) and balanced class weights (*class_weight* = ”balanced”). For one peptide (VSASPLLYTLIEK) negative values were present after external calibration. Those values were replaced by the minimal positive value of the respective peptide measured over all samples. Two peptides were removed (ASDTAMYYCAR and LVGGPMDASVEEEGVRR) as they were not reliably quantified leaving 48 peptides for the analysis. For every patient the first sample measured was selected (n = 165). All patients with unknown WHO grade/outcome were neglected. All data were log2-transformed and scaled to 0 mean and 1 variance fitted on the training data (*sklearn*.*preprocessing*.*StandardScaler*). The model was trained and validated using a shuffled stratified 5-fold cross-validation (*sklearn*.*model_selection*.*StratifiedKFold*) to assure that every split has a comparable case-to-control ratio and that every sample was used in 4 runs for training and in the remaining run for validating the trained model not including this sample. For reproducibility the seed was fixed to 42. For models trained on established risk assessments scores, only samples for which the respective score was determined were included in model construction and testing.

Decision function, ROC-Curve, accuracy, sensitivity and specificity were calculated using *scikit-learn 0*.*23*.*2*. For the Kaplan-Meier estimate *lifelines 0*.*26*.*0* [54] was used. The data were divided in death and survival predicted cases. For the dying patients, the days until death were included in the model. The samples for people who left the hospital alive were censored. Samples with missing time until outcome for patients who died were neglected. The confidence intervals were calculated using Greenwood’s Exponential formula as implemented in *lifelines 0*.*26*.*0* (*alpha* = 0.05).

In addition, a predictor based on the extra-trees algorithm implemented in *scikit-learn 0*.*23*.*2* (*sklearn*.*ensemble*.*ExtraTreesClassifier*) was evaluated. The same approach as described above was applied with the differences that the data weren’t log2-transformed and scaled as this isn’t needed for a tree-based classifier. In addition, the maximal depth of the trees was set to 3 (*max_depth* = 3) to compensate for overfitting issues due to limited data set size. Feature importances were extracted from a model trained on all data (n=165) without splitting the data set.

