## Supplementary Figures for "A multiplex protein panel assay determines disease severity and is prognostic about outcome in COVID-19 patients"

This file contains:
Supplementary Figures 1-4

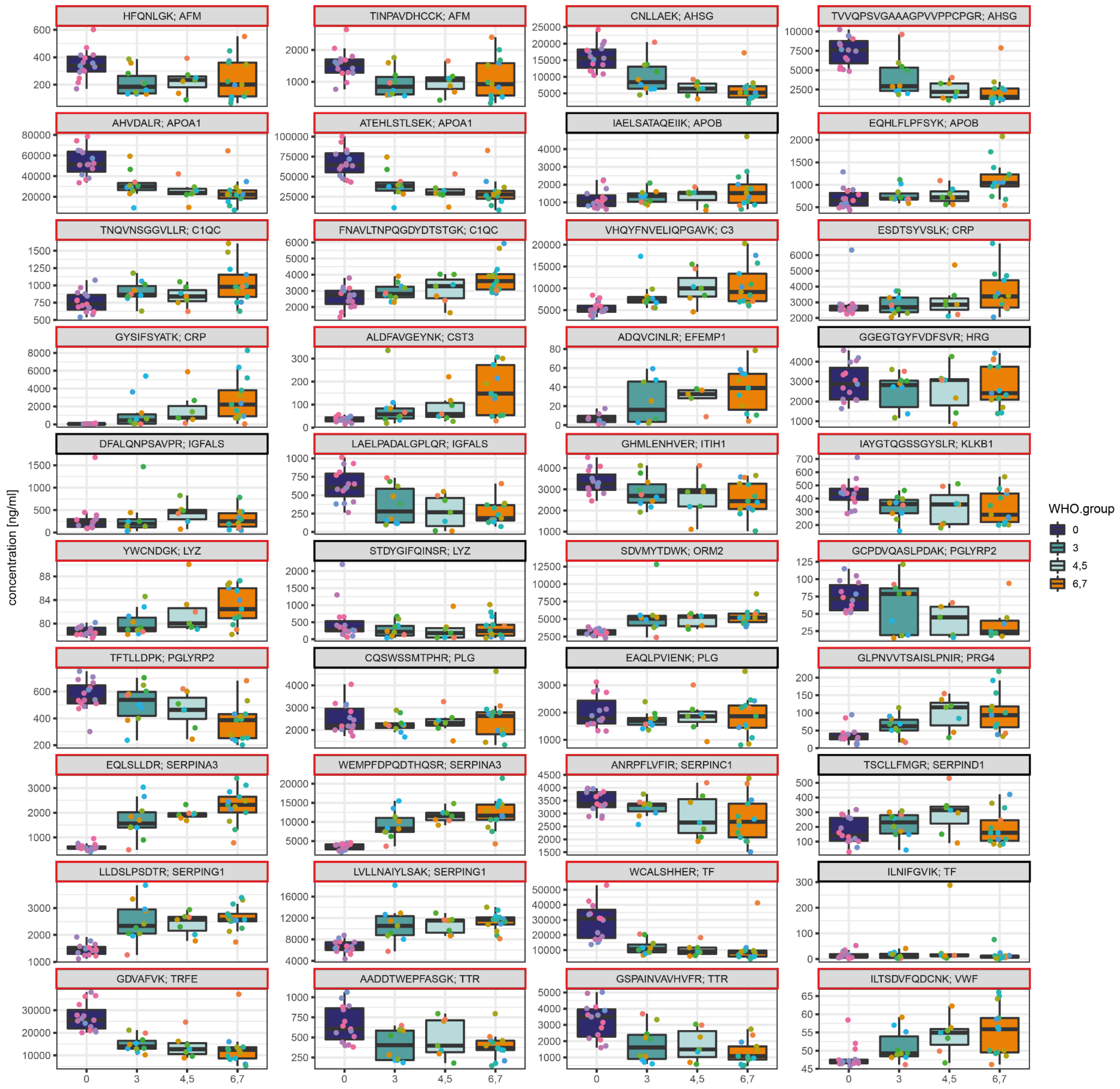


**Supplementary Figure 1. Abundance of selected peptides in different COVID-19 severity groups and healthy control samples.** Absolute concentration of all peptides quantified in cohort 1 (citrate plasma samples obtained during the 'first wave' in March 2020) plotted against the COVID-19 treatment escalation score. Note that the cohort also includes healthy COVID-19 negative control samples (blue; WHO score = 0). Peptides with a significant trend against COVID-19 severity as estimated on the respective WHO severity groups not-infected (0), mild (3), moderate (4 & 5) and severe (6 & 7) disease are highlighted in red ($P < 0.05$ after multiple testing correction). Data from n=15 (WHO 0), n=10 (WHO 3), n=4 (WHO 4), n=3 (WHO 5), n=3 (WHO 6), n=10 (WHO7).

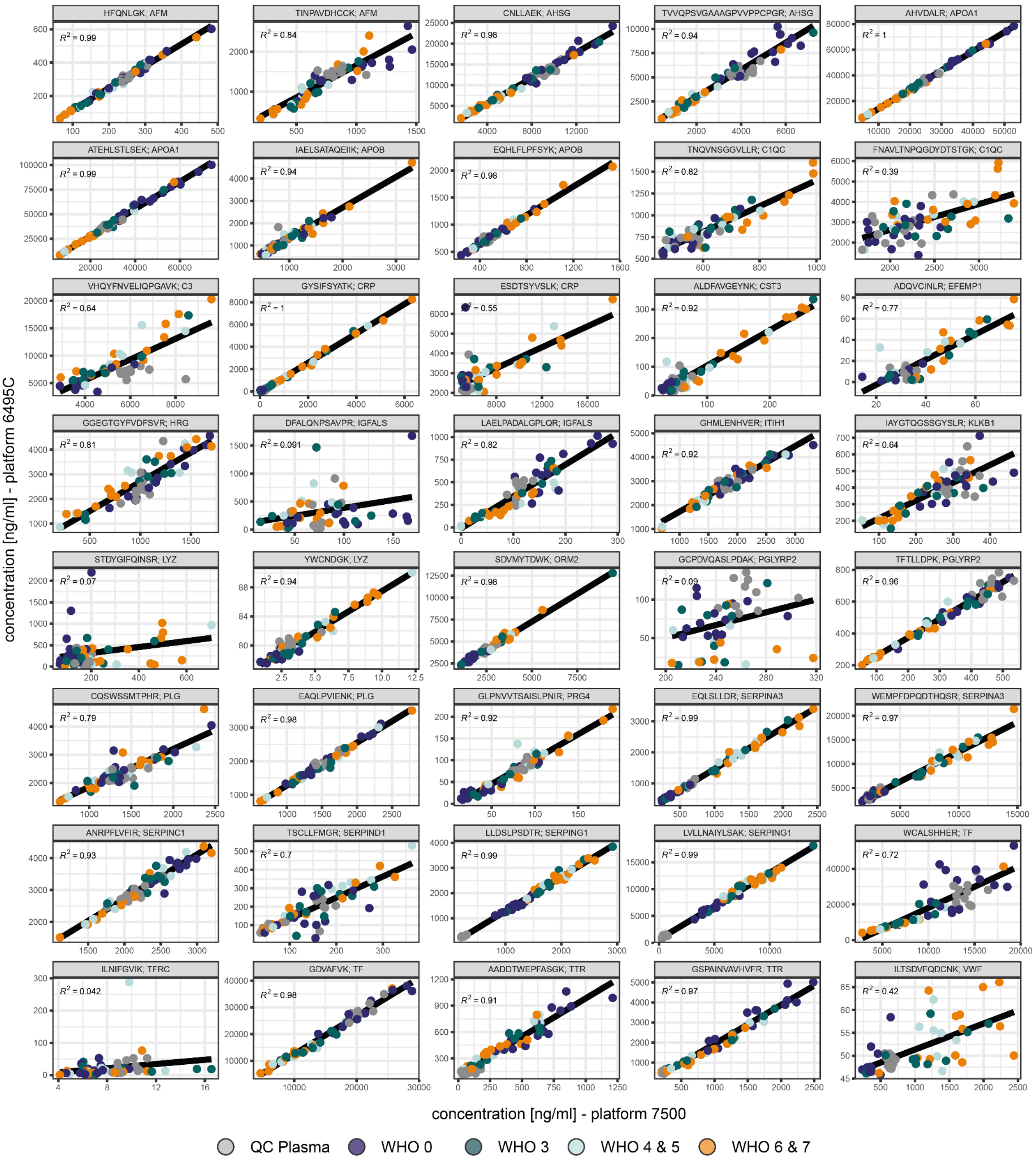


**Supplementary Figure 2. Comparison of the obtained absolute quantities of the putative peptide biomarkers in plasma samples measured on two LC-MRM platforms.** Samples were prepared, split in two aliquots, and measured on two different LC-MRM platforms, applying two sets of independently optimised MRM transitions, and operated in two laboratories. Shown are the linear correlations of the absolute concentration of all quantified peptides. Obtained R2 values of the linear fit as indicated. Values on the y-axis (platform 1) were measured on a 6495C (Agilent) instrument, values on y-axis (platform 2) were measured on a 7500 (SCIEX) instrument. All data based on n=15 (WHO 0), n=10 (WHO 3), n=4 (WHO 4), n=3 (WHO 5), n=3 (WHO 6), n=10 (WHO7) and n=12 (QC Plasma). Peptides are ordered alphabetically according to gene names. Peptides with poor correlation (R2 < 0.6) between both platforms are FNAVLTNPQGDYDTSTGK (C1QC), ESDTSYVSLK (CRP), DFALQNPSAVPR (IGFALS), STDYGIFQINSR (LYZ), GCPDVQASLPDAK (PGLYRP2), ILTSDVFQDCNK (VWF), ILNIFGVIK (TFRC). In the case of ESDTSYVSLK, this is largely due to one outlier (removal increases the R2 to 0.76). The other peptides are low abundant, increasing variance.

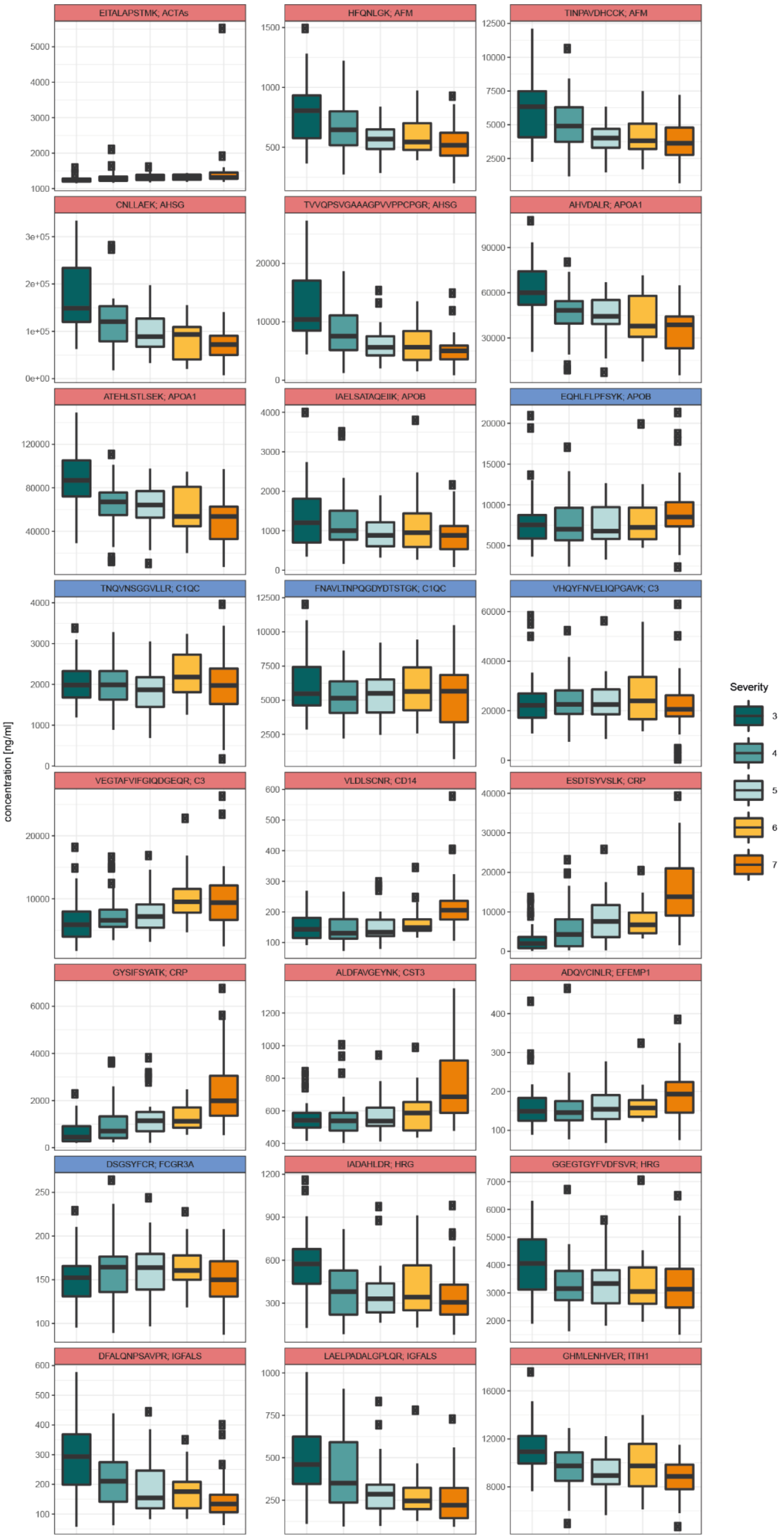
EITALAPSTMK; ACTAs
HFQNLGK; AFM
TINPAVDHCCK; AFM
CNLLAEK; AHSG
TVVQPSVGAAAGPVVPPCPGR; AHSG
AHVDALR; APOA1
ATEHLSTLSEK; APOA1
IAELSATAQEIIK; APOB
EQHLFLPFSYK; APOB
TNQVNSGGVLLR; C1QC
FNAVLTNPQGDYDTSTGK; C1QC
VHQYFNVELIQPGAVK; C3
VEGTAFVIFGIQDGEQR; C3
VLDLSCNR; CD14
ESDTSYVSLK; CRP
GYSIFSYATK; CRP
ALDFAVGEYNK; CST3
ADQVCINLR; EFEMP1
DSGSYFCR; FCGR3A
IADAHLDR; HRG
GGEGTGYFVDFSVR; HRG
DFALQNPSAVPR; IGFALS
LAELPADALGPLQR; IGFALS
GHMLENHVER; ITIH1
concentration [ng/ml]
Severity
3
4
5
6
7

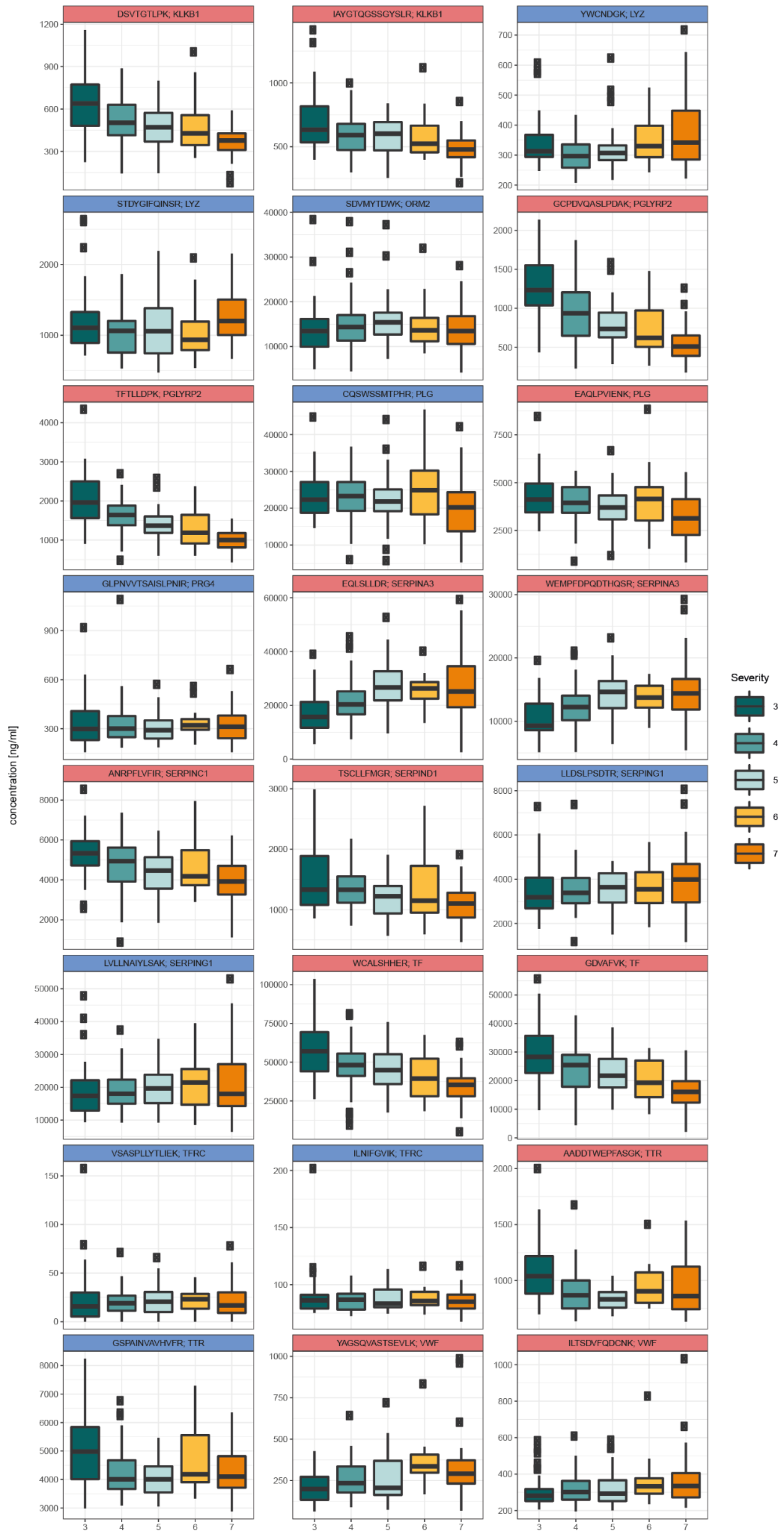


**Supplementary Figure 3. Abundance of selected peptides in different COVID-19 severity groups.** Absolute concentration of all peptides quantified in cohort 2 (plasma samples obtained during the second wave of the pandemic) plotted against the COVID-19 treatment escalation score. Peptides with a significant trend against COVID-19 severity as estimated on the respective WHO severity score are highlighted in red (Kendall´s Tau statistics, $P > 0.05$ after multiple testing correction). Data from the first time-point obtained for each individual; n=38 (WHO 3), n=45 (WHO 4), n=27 (WHO 5), n=16 (WHO 6), n=39 (WHO7). Peptides are ordered by their corresponding gene names.

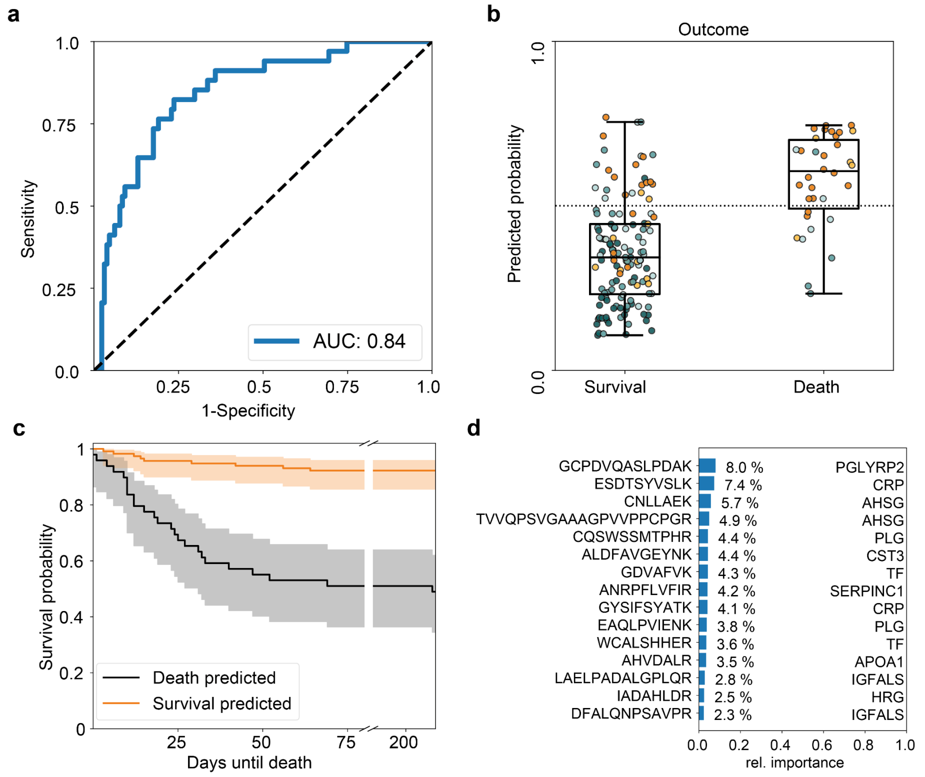


**Supplementary Figure 4. COVID19 progression prediction in a second wave COVID-19 cohort using an extra-trees model. a)** ROC-Curve for the prediction of death from the first time point measured for every patient using an extra-trees classifier. **b)** Boxplot of the predicted probability sorted according to the outcome and colored with respect to the WHO grade at the day the sample was taken. **c)** Kaplan-Meier estimate of the survival function for non-survival predicted cases (black) and survival predicted cases (orange) with confidence interval (alpha=0.05). All predictions were done on withheld samples that were not used for training. **d)** Feature importances for a model trained on all samples highlighting the corresponding protein/gene for each peptide. Notably, for several proteins (CRP, AHSG, PLG, TF, IGFALS) more than one peptide was ranked as an important feature (Top 15 features) by the predictor.
